# Neural bases of Frustration-Aggression Theory: A multi-domain meta-analysis of functional neuroimaging studies

**DOI:** 10.1101/2021.05.12.21257119

**Authors:** Jules R. Dugré, Stéphane Potvin

## Abstract

Early evidence suggests that unexpected non-reward may increase the risk for aggressive behaviors. Despite the growing interest in understanding brain functions that may be implicated in aggressive behaviors, the neural processes underlying such frustrative events remain largely unknown. Furthermore, meta-analytic results have produced discrepant results, potentially due to substantial differences in the definition of anger/aggression constructs. Therefore, coordinate-based meta-analyses on unexpected non-reward and retaliatory behaviors in healthy subjects were conducted. Conjunction analyses were further examined to discover overlapping brain activations across these meta-analytical maps. Frustrative non-reward deactivated the orbitofrontal cortex, ventral striatum and posterior cingulate cortex, whereas increased activations were observed in midcingulo-insular regions, as well as dorsomedial prefrontal cortex, amygdala, thalamus and periaqueductal gray, when using liberal threshold. Retaliation activated of midcingulo-insular regions, the dorsal caudate and the primary somatosensory cortex. Conjunction analyses revealed that both strongly activated midcingulo-insular regions. Our results underscore the role of anterior midcingulate/pre-supplementary motor area and fronto-insular cortex in both frustration and retaliatory behaviors. A neurobiological framework for understanding frustration-based impulsive aggression is provided.

## 1. Introduction

Nearly a century ago, Dollard and colleagues proposed that some forms of aggression would appear in an inhibiting context, during which an object would interfere with goal attainment (Dollard, Miller, Doob, Mowrer, & Sears, 1939), therefore enhancing the risk for aggressive behaviors. In the following years, several researchers have criticized the mutually exclusive relationship between frustration^1^ and aggression, suggesting that frustration may lead to a general negative emotional state (e.g., neuroticism, sadness, anger) and not directly to aggression (Berkowitz, 1989). More precisely, it has been argued that such inhibiting events will lead to aggression if the former context causes sufficient negative affect to enable aggressive inclinations (Berkowitz, 1989). Despite the fact that it has been almost forgotten in current research, the Frustration-Aggression theory remains one of the most prominent theory of aggression.

Reward processing and loss aversion are inherent aspects of human motivation. Neurobiological research has shown a strong implication of ventral striatum, medial OFC/vmPFC and PCC/Precuneus in reward processes (Bartra, McGuire, & Kable, 2013; Liu, Hairston, Schrier, & Fan, 2011; Oldham et al., 2018), whereas punishment/loss processing is known to recruit anterior insula/vlPFC and lateral OFC (Bartra et al., 2013; Dugré, Dumais, Bitar, & Potvin, 2018; Liu et al., 2011; Oldham et al., 2018). Interestingly, receiving offers from someone who follows (or not) social norms may involve reward/punishment processes (7). Indeed, receiving fair monetary offers seems to recruit similar brain regions to those involved in reward processing (e.g., medial OFC/vmPFC, PCC/Precuneus) while receiving unfair monetary offers seems to recruit regions involved in punishment processng (aINS/vlPFC & lateral OFC), respectively (Feng, Luo, & Krueger, 2015). In fact, norm violations share several features with frustration. Indeed, given that we implicitly expect that others will follow social norms (e.g., fairness/reward) (Bicchieri & Chavez, 2010), norm violations (e.g., unfairness/frustration) act as a barrier to goal-directed behaviors. Additionally, both unexpected non-reward and norm violations elicit negative emotional responses. For instance, past researchers have found that receiving unfair monetary offers induce negative affect such as anger, sadness and feelings of being betrayed (Fatfouta, Meshi, Merkl, & Heekeren, 2018; Gradin et al., 2015; Paz et al., 2017). Furthermore, it has been shown that negative emotional arousal (e.g., sadness, anger & heightened skin conductance) further increases the risk of rejecting unfair offers (Harlé, Chang, van’t Wout, & Sanfey, 2012; Harlé & Sanfey, 2007; C. Liu, Chai, & Yu, 2016; Osumi & Ohira, 2010; van’t Wout, Chang, & Sanfey, 2010). On neurobiological grounds, it has also been observed that the activation of the anterior insula (during unfair offers) may mediate the relationship between negative emotions (i.e. sadness) and rejection rates (Harlé et al., 2012). Thus, in line with Berkowitz assumptions (1989), these results suggest that propensity towards retaliatory behaviors may be increased when the frustrative event give rises to heightened negative emotional responses. However, in the neuroscientific literature, the neural mechanisms elicited during frustrative events are largely unknown. In fact, frustration processing (or non-reward processing) has often been blurred by the dichotomous reward-punishment categorization and its overutilization as a baseline condition in fMRI task contrasts. Nonetheless, it may hide a particular series of actions involving the interaction between both reward and punishment processes.

In the last decade, frustration has been thought to be implicated in several psychopathologies that are at risk for aggressive behaviors (e.g., irritability, CD/ASPD, see (Bertsch, Florange, & Herpertz, 2020; Blair, 2010b; Harenski & Kiehl, 2010; Leibenluft, 2017)). Therefore, uncovering the neural bases of frustration processing is of great importance for our understanding of processes by which aggressive behaviors may likely arise. However, past meta-analyses did not produce reliable neurobiological markers of aggressive behaviors, potentially due to a substantial heterogeneity in the definition of aggression-related emotion and behavioral responses. For instance, a meta-analysis of fMRI studies on “*state anger*” (Puiu et al., 2020), which included a wide range of fMRI task contrasts (e.g., punishing others, rejecting unfair offers, retaliatory behaviors and passively reacting to provocation), revealed significant activations in the mPFC, pregenual ACC and right anterior insula. Similarly, another meta-analysis was conducted on “*trait aggression*” which included a variety of different tasks (e.g., theory of mind, passive viewing, working memory, probabilistic reversal task) with distinct psychiatric populations (e.g., schizophrenia, intermittent explosive disorder, antisocial personality disorder) and showed that only the precuneus emerged as significant cluster (Wong et al., 2019). The authors carried out an additional meta-analysis on “*elicited aggression*”, which was more precisely defined (e.g., Taylor aggression paradigm and Violent first-person shooter games), and the meta-analysis yielded activations in the postcentral gyrus (Wong et al., 2019). Finally, in a recent meta-analysis on “*anger experience*”, the authors have included heterogeneous fMRI task domains including retaliatory behaviors, social exclusion and anger imagery (Sorella, Grecucci, Piretti, & Job, 2021), revealing activations in bilateral insula/vlPFC. Despite that these major differences in definition may partially explain the discrepancies in brain regions, it nonetheless illustrates the importance of dissecting homogeneous constructs using theory-driven terminology.

Based on a literature review, Blair (2016) has suggested that impulsive aggression may involve interactions between acute threat response (i.e., amygdala, hypothalamus and periaqueductal gray [PAG]), the vmPFC and the mid-cingulo-insular network (i.e., dACC, aMCC/pre-SMA and bilateral fronto-insular cortex). The current study therefore aims to unveilthe neural bases of the frustration-aggression theory and provide meta-analytical support for a neurobiological model of frustration-based impulsive aggression. More precisely, we sought to examine to what extent the frustrative non-reward and retaliatory behaviors spatially overlapped altogether, in order to provide a model of how this series of actions operate at the neurobiological level.

## 2. Methods

### 2.1. Data Inclusion

In the current meta-analysis, the search strategies were the BrainMap Database as well as the reference lists of recent meta-analyses (See Figure 1). Inclusion criteria were the use of whole-brain analyses in healthy controls reported in a standard reference space (Talairach/Tournoux, Montreal Neurological Institute [MNI]) using functional Magnetic Resonance Imaging (fMRI) or Positron Emission Tomography. Talairach coordinates were converted into MNI space before using them in analyses.

**Figure 1.**
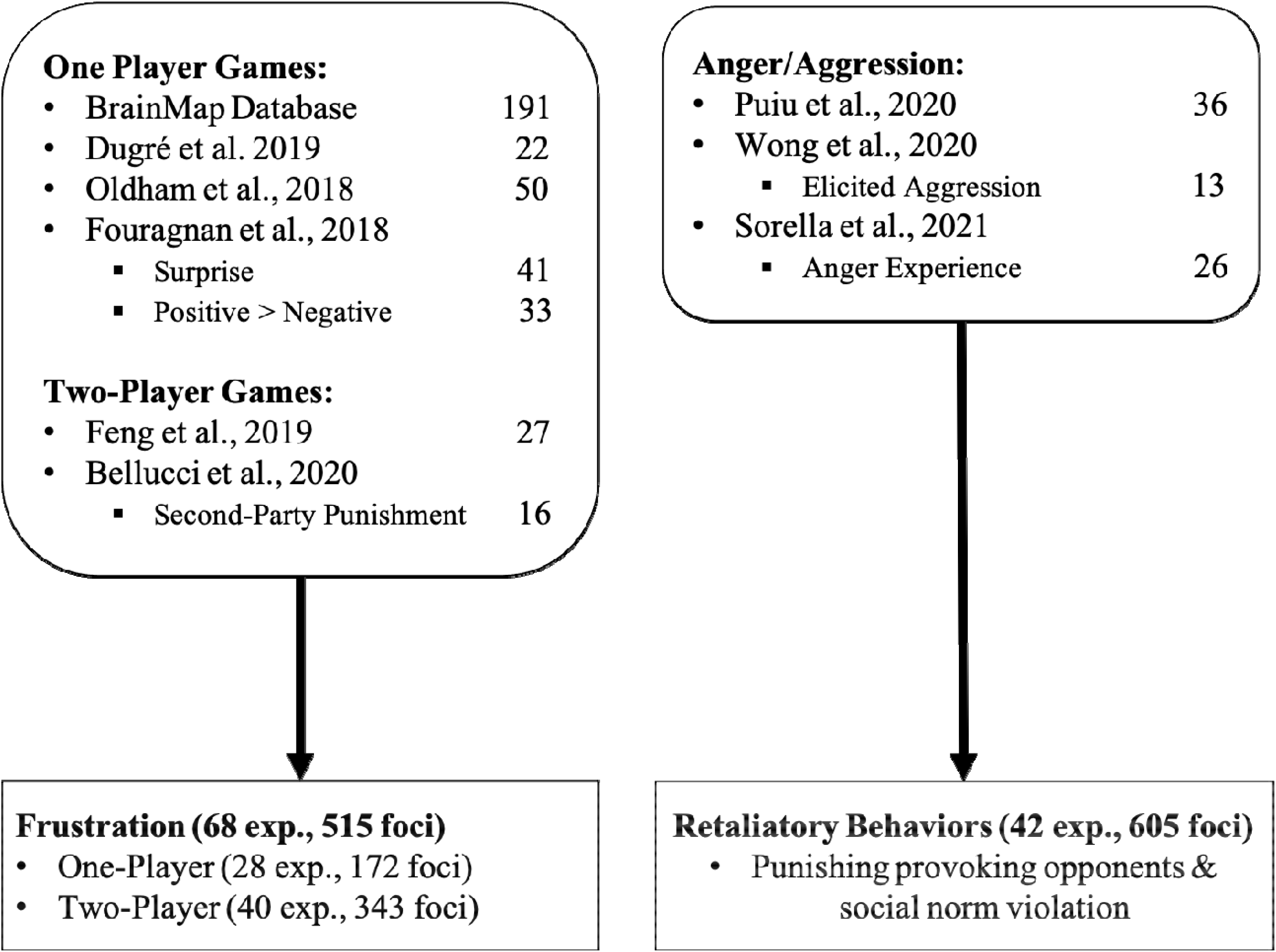
Flowchart of the literature search for Frustration and Retaliatory Behaviors.

Frustrative events were defined as the withholding or the removal of an expect reward by others that was not performance-contingent (e.g., rewarded response inhibition tasks). Experiments involving frustrative events were retrieved by screening the BrainMap Database (i.e., *Reward Paradigm Class* and *Normal Mapping Context* as search terms which allowed to retrieve191 papers). Another search was done by screening references from recent meta-analyses on reward/punishment processing (Dugré et al., 2018; Fouragnan, Retzler, & Philiastides, 2018; Oldham et al., 2018). For the two-player games, we extracted data from the reference lists of recent meta-analyses on the Ultimatum Game (Bellucci, Camilleri, Iyengar, Eickhoff, & Krueger, 2020; Feng et al., 2019). For the frustration meta-analysis, we pooled data from the one-player (e.g., Non-Reward Outcome in the Reward trial of the Monetary Incentive Delay Task) and two-player games (i.e., Unfair condition of the Ultimatum game), resulting in (Frustration_SINGLE_ + Unfair_MULTI_) versus (Reward_SINGLE_ + Fair_MULTI_). Indeed, this was done since frustration and unfairness shares a number of similarities. For instance, receiving a fair offer from the proposer results in a reward for both parties, in the Ultimatum Game. However, since the receiver implicitly expects the proposer to follow social norms about fairness (Bicchieri & Chavez, 2010), violating this norm yields a non-rewarding outcome for the receiver (i.e., frustrative context).

Finally, the search for experiments on retaliatory behaviors was done by screening the reference lists of recent meta-analyses on aggressivity/anger (Puiu et al., 2020; Sorella et al., 2021; Wong et al., 2019). Experiments on retaliatory behaviors were defined by an explicit decision to punish another player that had provoked the participant (e.g., removing points, delivering a shock/noise).

### 2.2. Activation Likelihood Estimate method

Experiments’ coordinates were used for spatial convergence using the Activation Likelihood Estimate method (GingerALE version 3.0.2, (Eickhoff, Bzdok, Laird, Kurth, & Fox, 2012; Eickhoff et al., 2009); http://www.brainmap.org/ale/). For each experiment, a 3D gaussian probability distribution was modelled around each coordinate foci, weighted by the number of subjects in the experiment. This method is performed to account for spatial uncertainty due to template and between-subject variance (Eickhoff et al., 2012; Eickhoff et al., 2009). It also ensures that multiple coordinates from a single experiment does not jointly influence the modeled activation value of a single voxel. The probabilities of all activation foci in an experiment were then combined. Voxel-wise ALE scores arise from the union across all these experiment maps. Consequently, a cluster-level corrected threshold was applied on the voxel-wise image. The size of the supra-threshold clusters was compared against a null distribution of cluster sizes derived from simulation of datasets. We used the following statistical threshold: p<0.001 at voxel-level and FWE-p<0.05 at a cluster-level with 5000 permutations. Results were also reported using a more liberal statistical threshold of p<0.001 uncorrected given that activation of subcortical regions during tasks eliciting emotional response may be suppressed by regulatory mechanisms in healthy subjects. Indeed, as hypothesized by Blair (2016), subcortical and midbrain regions including the amygdala, hypothalamus and PAG would be observed.

Conjunction analyses were carried out to examine spatial overlap between the meta-analytical maps. Here, test for convergence was determined by the intersection between the meta-analyses. These results are reported for a p<0.0001 using 10,000 permutations.

## 3. Results

### 3.1. Main Effects of Frustration-Aggression Phases

#### 3.1.1. Frustration Processing

The meta-analysis of main effect of frustration (non-reward) comprised 68 experiments (515 foci, 1801 subjects). ALE meta-analysis revealed significant activations in bilateral fronto-insular cortex (FIC) and dACC/aMCC & pre-SMA (see Table 2; Figure 2). Furthermore, when using a more liberal statistical threshold (p<0.001 uncorrected), we observed activation in the dmPFC, medial dorsal nucleus of the thalamus, the amygdala and the periaqueductal grey (PAG) (see Table 2; Figure 2A). Contrasting Frustration_SINGLE_ versus Unfair_MULTI_ showed differences only in a small cluster of the pre-SMA (See Supplementary Material)

**Figure 2.**
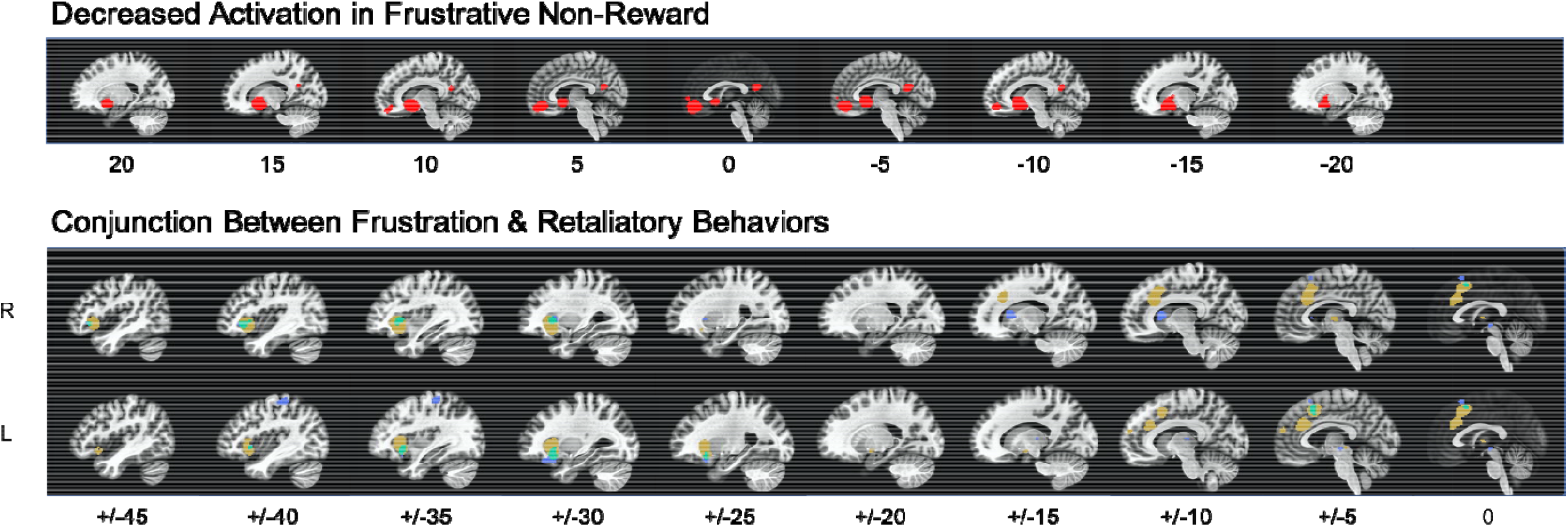
Main results of the meta-analysis on frustration and retaliatory behaviors. Red = Decreased activation in the meta-analysis on frustrative non-reward. Yellow = Increased activation in the meta-analysis on frustrative non-reward. Blue = Increased activation in the meta-analysis on retaliation. Green = results from the conjunction analysis between frustrative non-reward and retaliation.

Additionally, the reverse contrast examining the decreased activations after being frustrated (63 experiments, 395 foci and 3047 subjects) showed significant peak convergence in the ventral striatum (bilateral), medial OFC/vmPFC and the PCC/Precuneus (see Table 1; Figure 2). Contrasting Reward_SINGLE >_ Frustration_SINGLE_ versus Fair_MULTI_ showed differences in a small cluster encompassing the right putamen (See Supplementary Material).

**Table 1.** Main Effect of Frustration and Retaliatory Behaviors

| Main Peaks | MNI Coordinates |  |  | ALE | Voxels |
| --- | --- | --- | --- | --- | --- |
|  | X | Y | Z |  |  |
| ***Frustration*<sub>INCREASED</sub>** |  |  |  |  |  |
| dACC/aMCC/pre-SMA | 8 | 24 | 36 | 0.0334 | 1569 |
| FIC | 36 | 22 | -4 | 0.0528 | 1051 |
| FIC | -30 | 22 | 0 | 0.0499 | 1000 |
| dmPFC† | -8 | 60 | 20 | 0.0279 | 54 |
| Thalamus (MD) † | 4 | -12 | 4 | 0.0212 | 47 |
| Amygdala† | -18 | -4 | -10 | 0.0197 | 24 |
| PAG† | -4 | -28 | -4 | 0.0205 | 21 |
| ***Frustration*<sub>DECREASED</sub>** |  |  |  |  |  |
| vStriatum | 12 | 10 | -8 | 0.0455 | 2144 |
| vStriatum | -10 | 10 | -8 | 0.0357 | - |
| mOFC/vmPFC | 2 | 46 | -12 | 0.0239 | 940 |
| PCC/Precuneus | -6 | -58 | 18 | 0.0158 | 451 |
| ***Retaliatory Behaviors*** |  |  |  |  |  |
| FIC | 30 | 20 | 2 | 0.0341 | 288 |
| FIC | -30 | 18 | -14 | 0.0365 | 274 |
| dStriatum | 12 | 14 | 10 | 0.0293 | 159 |
| postcentral; IPL | -38 | -32 | 62 | 0.0251 | 126 |
| aMCC; pre-SMA; SMA | -4 | 16 | 52 | 0.0242 | 121 |
| PAG† | -4 | -20 | -6 | 0.0246 | 44 |
| Thalamus (VPL)† | -14 | -22 | 8 | 0.0195 | 23 |

*Note.* † = Peaks that emerged when using a more liberal statistical threshold ( $p < 0.001$ uncorrected). All other peaks are reported at $p < 0.001$ voxel and $cFWE < 0.05$ .

#### 3.1.2. Retaliatory Behaviors

Forty-two neuroimaging experiments were included in the ALE meta-analysis on retaliation (605 foci, 1285 subjects). Retaliatory behaviors were associated with activations in bilateral FIC, the dorsal striatum (i.e., caudate), the left postcentral gyrus/Inferior Parietal Lobule and the aMCC/pre-SMA (see Table 2; Figure 2). Using a more liberal threshold (p<0.001 uncorrected), additional cluster in the PAG and the ventral posterolateral thalamus were observed.

### 3.2. Conjunction Analyses

Conjunction analyses comparing Frustration_INCREASED_ and Retaliatory behaviors yielded overlap in bilateral FIC (Left: x=-32, y=20, z=-8, ALE=0.0255, 224 voxels; Right: x=32, y=20, z=2, ALE=0.0312, 204 voxels) as well as in the aMCC/pre-SMA (x=-4, y=16, z=50, ALE=0.0255, 98 voxels) (See Figure 2). No significant spatial overlap was observed between the Frustration_DECREASED_ and Retaliatory behaviors.

## 4. Discussion

The current study aimed to examine, through a meta-analysis, the neural bases of the core components of the frustration-aggression theory. Using a theory-driven definition of aggression-related constructs, this study also aimed to clarify the discrepancies in recent meta-analyses on anger/aggression and focus on uncovering processes which may increase the risk for reactive aggression. For instance, we observed that frustrative events yield decreased activations in the ventral striatum, medial OFC and PCC/precuneus while they activate bilateral fronto-insular regions (vlPFC, aINS, lOFC) and aMCC/pre-SMA. We also observed that frustration was associated with increased activations of the dmPFC, thalamus, amygdala and PAG when using a more liberal statistical threshold. Additionally, retaliatory behaviors were associated with bilateral FIC, aMCC/pre-SMA as well as dorsal striatum and the primary somatosensory cortex (S1) as well as the PAG and thalamus only when using a more liberal threshold. Finally, conjunction analyses revealed that bilateral FIC may be common to frustration context and retaliatory behaviors. Results from this meta-analysis are of great significance as they provide empirical support for Blair’s theoretical framework of impulsive aggression (Blair, 2016), emphasizing the primary role of vmPFC, dACC/aMCC (referred to dmPFC in Blair, (2016), anterior insula, amygdala and PAG in reactive aggression. We also observed that the Frustration-Aggression network may involve other brain regions that have been neglected thus far, such as the thalamus, caudate nucleus and SI. More importantly, our results provide additional insights about the potential mechanism underlying frustration-based impulsive aggression (See Figure 3).

**Figure 3.**
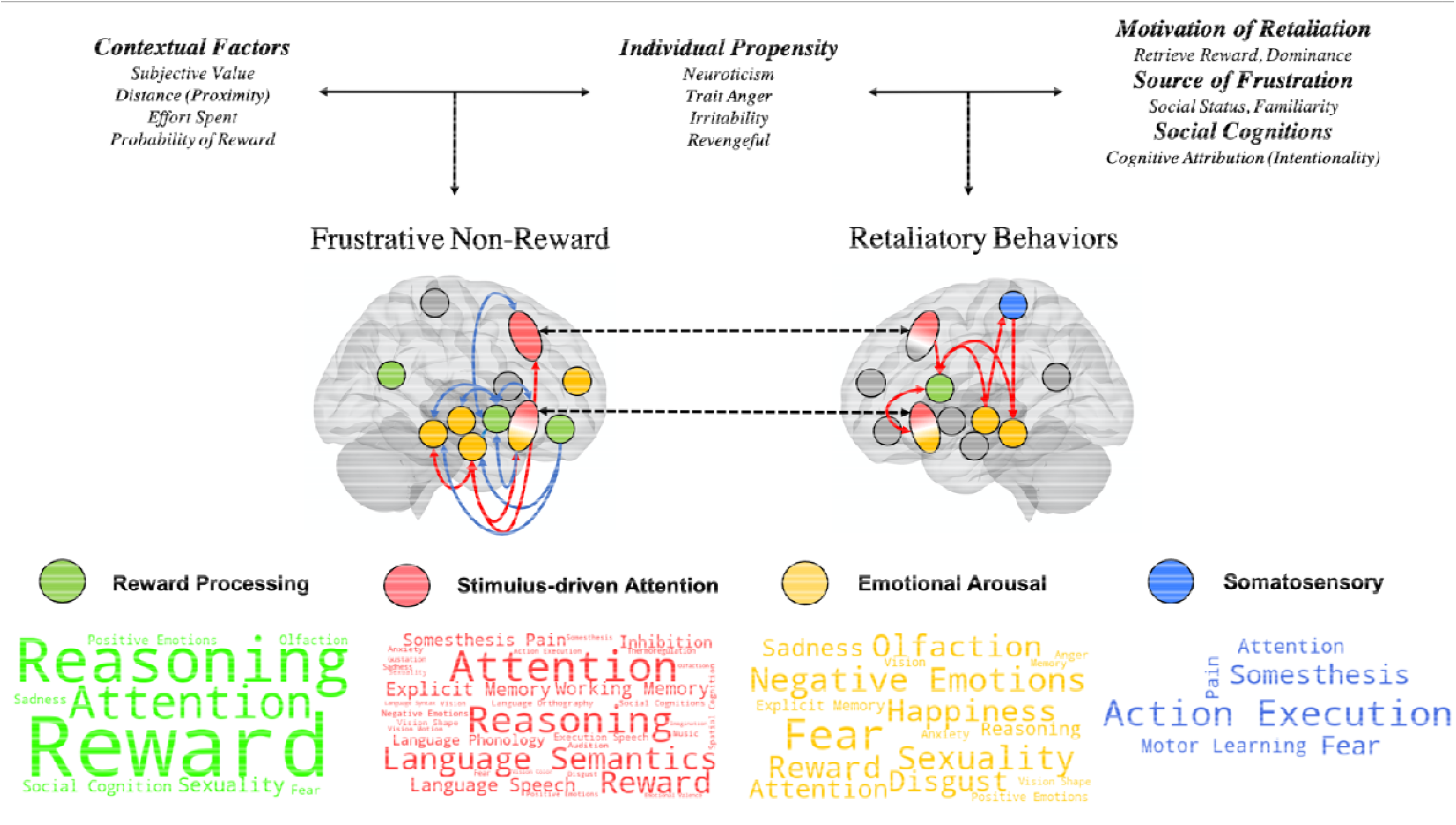
Summary of the neurobiological model of Frustration-based impulsive aggression. Lines represent interaction between subsystems as supported by literature. Blue line = negative relationship between brain regions; Red lines = positive relationship between brain regions. Black dotted line = brain regions that significantly overlapped between frustrative non-reward and retaliatory behaviors. Word Cloud were produced using the Behavioral Analysis Plugin of the Multi-image Analysis GUI which utilize the BrainMap database (Lancaster et al., 2012). Bigger font represents higher Z-score. Every word presented in the Figure showed a statistically significant Z-Score (higher than 3.0) with Bonferroni correction for multiple comparison (p<0.05).

### 4.1. Integrative Neurobiological Model of Frustration-Aggression sequence

Responding adequately to frustrative events is thought to be an evolutionary function which is necessary for species survival. For instance, aggressive behaviors may be appropriate to defend, protect or punish others from stealing community’s food. Hence, aggressive behaviors which resulted from the omission of an expected reward have been observed in a variety of species such as in rodents (Burokas, Gutiérrez□Cuesta, Martín□García, & Maldonado, 2012; de Almeida & Miczek, 2002; Gallup, 1965), pigs (Arnone & Dantzer, 1980; Dantzer, Arnone, & Mormede, 1980), fishes (Vindas et al., 2012; Vindas et al., 2014) as well as in birds (Azrin, Hutchinson, & Hake, 1966; Duncan & Wood-Gush, 1971). To our knowledge, the current meta-analysis is the first to specifically examine the neural bases of frustrative-aggression sequence. In this meta-analysis, we followed Berkowitz’ (1989) proposition suggesting that withholding/removing an expected reward would enhance emotional arousal, which further increase the risk for reactive aggression. Given that several psychopathologies may exhibit enhanced risk for frustration and reactive aggression (Bertsch et al., 2020; Leibenluft, 2017), its implication for clinical population are also discussed.

#### 4.1.1. The Activation and Deactivation systems underlying Frustration

Evidence from the current meta-analysis suggests that two processes may co-occur when a reward has been frustrated. First, we found that withholding an expected reward results in significant decreased activations of the VS, vmPFC/mOFC and the vPCC. Interestingly, research has consistently shown that the ventral striatum (Abler, Erk, & Walter, 2007; Abler, Walter, & Erk, 2005; Bjork, Chen, Smith, & Hommer, 2010; Bjork, Smith, & Hommer, 2008; V. B. Gradin et al., 2013; McClure, Berns, & Montague, 2003; Pessiglione, Seymour, Flandin, Dolan, & Frith, 2006) and the vmPFC/mOFC (Bjork et al., 2004; Bjork et al., 2008; Knutson, Fong, Adams, Varner, & Hommer, 2001; Knutson, Fong, Bennett, Adams, & Hommer, 2003) actually deactivate in frustrative events. In addition, evidence functional and structural MRI studies has shown strong connectivity between the vmPFC and VS (Cauda et al., 2011; Di Martino et al., 2008; Hart, Leung, & Balleine, 2014) which subserve goal-directed learning (Hart et al., 2014), supporting their frequent co-activation in meta-analyses on reward processing (Bartra et al., 2013; Liu et al., 2011; Oldham et al., 2018). However, results from past meta-analyses suggest that activation vPCC may be specific to reward processing (Bartra et al., 2013; Liu et al., 2011; Oldham et al., 2018), and may not be activated when receiving punishment (Dugré et al., 2018). Hence, there is currently no evidence of vPCC deactivation during unexpected non-reward, indicating potential artefactual results due to task-contrast (reward > unexpected non-reward).

Besides this deactivation system, we also found increased activation in bilateral anterior FIC as well as the dACC/aMCC and pre-SMA. Indeed, functional characterization analyses revealed their strong relationship with attention, reasoning, reward, memory, inhibition and pain. These brain regions are crucial nodes of the midcingulo-insular network (Uddin, Yeo, & Spreng, 2019) which is mainly involved in detection of salient information in the environment (i.e., stimulus-driven attention). The co-occurrence of the aMCC/pre-SMA and anterior FIC has also been noticed a wide range of regulatory mechanisms such as conflict monitoring and emotion regulation strategies (Buhle et al., 2014; Morawetz, Bode, Derntl, & Heekeren, 2017) and response to surprising salient outcome (Fouragnan et al., 2018). More specifically, it has been suggested that the anterior insula may play a major role in emotional awareness by integrating interoceptive prediction errors with and top-down signals originating from the aMCC/pre-SMA and PFC (Gu, Hof, Friston, & Fan, 2013). Hence, findings from our meta-analysis highlight the role of the midcingulo-insular regions in frustrative non-reward.

Moreover, we observed significant activation of the dmPFC, medial dorsal thalamus, amygdala and PAG when using a more liberal statistical threshold (p<0.001 uncorrected). These brain regions were further characterized as being involved in negative emotions (i.e., fear, sadness, anger, disgust), attention, explicit memory but also in happiness and reward. Relationships between these cortical (i.e., aMCC/pre-SMA, FIC and dmPFC) and subcortical structures (i.e., amygdala, thalamus, PAG) has often been reported in threat conditioning (Fullana et al., 2016; Mechias, Etkin, & Kalisch, 2010), physiological arousal ((Hartley & Phelps, 2010) for meta-analysis see (Beissner, Meissner, Bär, & Napadow, 2013). The amygdala is involved in affective-motivational component of pain processing, while the PAG is known for playing a key role endogenous pain modulation and stress-induced analgesia (Ong, Stohler, & Herr, 2019; Tanasescu, Cottam, Condon, Tench, & Auer, 2016). In addition, past meta-analyses and studies suggested that the PAG may play a non-negligeable role in processing negative affect given its activation, not only in pain conditions, but also when viewing negative emotional pictures (Buhle et al., 2013; Kober et al., 2008; Wager et al., 2008). Hence, as proposed by Blair (Blair, 2016), these brain regions (acute threat system) may play a crucial role in heightening emotional responsiveness to the frustrative event, further mediating impulsive aggression.

Neurobiological markers of frustrative events may thus be characterized by interaction between these 3 subsystems. Evidence from structural and functional connectivity studies suggests that the ventral part of the anterior insula demonstrate close connections with the VS, amygdala and hypothalamus and dmPFC (to a lesser extent), whereas the dorsal part is rather connected with frontal pole, dACC and the aMCC/pre-SMA, which are subserving cognitive control processes (Deen, Pitskel, & Pelphrey, 2011; Ghaziri et al., 2018; Menon et al., 2020; Nomi et al., 2016). Indeed, the IMAGEN Consortium (n=1510) has shown that when rewards were unexpectedly omitted, the ventral striatum was negatively correlated with activity of the insular cortex (bilaterally), the aMCC/pre-SMA and the thalamus (Cao et al., 2019). Interestingly, similar negative relationship has also been observed in pain studies showing deactivation of the VS (Aharon, Becerraa, Chabris, & Borsooka, 2006; Becerra & Borsook, 2008; Becerra, Navratilova, Porreca, & Borsook, 2013) and increased activation of aMCC/pre-SMA and insular cortex during pain onset (Meta-analyses: (Duerden & Albanese, 2013; Friebel, Eickhoff, & Lotze, 2011; Jensen et al., 2016; Lanz, Seifert, & Maihöfner, 2011; Tanasescu et al., 2016; Xu et al., 2020). The negative associations are consistent with the pattern of activations and de-activations observed in the current meta-analysis. Considering these, the ventral aINS may thus receive VS signals, enabling the threat system through its projection to the amygdala and hypothalamus when perceiving non-reward as a personal threat. Co-occurrently, projections from the dorsal part of the aINS to the aMCC/pre-SMA may reflect attentional shift towards the source of blockage (salient stimuli).

Furthermore, deactivation of the vmPFC may also play a prominent role in frustrative non-reward. Indeed, it has been shown that the vmPFC has a multifaceted role in emotional responding given its connectivity with both VS (reward) and Amygdala (threat) (Haber, 2016; Haber & Knutson, 2010; Hiser & Koenigs, 2018; Schneider & Koenigs, 2017). Indeed, vmPFC has been linked with subjective valuation and probability of reward (Bartra et al., 2013). Lesions studies have shown that the vmPFC is crucial for reinforcement-based decision making (Fellows & Farah, 2007; Koenigs, Kruepke, & Newman, 2010; Schneider & Koenigs, 2017) and is associated with increased appetite for risky decision-making (hot decisions)(Clark et al., 2008; Spaniol, Di Muro, & Ciaramelli, 2019; Studer, Manes, Humphreys, Robbins, & Clark, 2015). Thus, blocking a subjectively salient reward may deactivate the vmPFC, and alter subsequent decision-making in affect-rich contexts. This concurs with frustration theories proposing that the subjective value of rewards may explain interindividual difference in emotional arousal after a frustrating event (Berkowitz, 1989; Breuer & Elson). For instance, participants receiving anodal stimulation of the vmPFC perceived unfair offers as fairer than those in the sham group (Gilam et al., 2018), which indicates that stimulation of the vmPFC may modulate subjective value toward a given stimulus and further decrease the need for regulating (dACC/FIC) brain regions involved in the acute threat system (amygdala, thalamus, PAG). The vmPFC is also thought to play a role in generating and suppressing affect by a top-down regulating process in which the vmPFC may serves as a brake to inhibit the amygdala and the PAG and adapt behavior when evaluating imminence of the threat (Hiser & Koenigs, 2018; Kim et al., 2011; Mobbs et al., 2007). Nevertheless, frustrative non-reward may be characterized by deactivation of the vmPFC and VS, which enable stimulus-driven attention (aMCC/pre-SMA and FIC) and disinhibit the acute threat system, induces vigilance and physiological arousal.

#### 4.1.2. Neural bases of Retaliatory Behaviors

In the past decades, researchers have attempted to model the core neural features of reactive aggression. For instance, it has been argued that the medial hypothalamus, amygdala and the PAG are crucial nodes for understanding retaliatory behaviors (Bertsch et al., 2020; Blair, 2016; Crowe & Blair, 2008; Gregg & Siegel, 2001; Lickley & Sebastian, 2018; Panksepp, 2004; Panksepp & Zellner, 2004). In our meta-analysis we found that retaliatory behaviors were mainly characterized by activation in the dorsal striatum and primary somatosensory cortex (SI, BA3 encompassing the hand network of the left hemisphere), as well as bilateral FIC and aMCC/pre-SMA. Through projections from these regions to the caudate nucleus ((Haber, 2016; Haber & Knutson, 2010; Leh, Ptito, Chakravarty, & Strafella, 2007; Xiaojin Liu et al., 2021), behavioral responses to frustration are likely to arise. Indeed, the dorsal striatum has been implicated in several goal-directed decision making and action selection and initiation (Balleine, Delgado, & Hikosaka, 2007; Haber & Knutson, 2010; O’Doherty et al., 2004). Additionally, lesions to dorsal striatum produce deficient defensive behaviors in rats (Kirkby & Kimble, 1968; Winocur, 1974), supporting the role of action preparation and motor control in fight-or-flight response. Thus, this result is somewhat expected given that paradigms used in humans to investigate retaliatory behaviors often involve decisions whereby participants select the severity of the punishment directed towards others. However, some authors suggests that the VS may be implicated in retaliatory behaviors through a mechanism involving pleasure to retaliate (Chester & DeWall, 2016, 2018). In this meta-analysis, we did not find any spatial convergence in VS and other brain regions involved in the reward system, indicating that retaliatory behaviors in healthy subjects may not be as “*pleasurable*” as previously expected. Nevertheless, we cannot rule out the possibility that the VS activation during retaliation (or when viewing others in pain) may be specific to some clinical populations such as CP/ASPD with psychopathic traits and/or high levels of sadistic traits (Decety, Chen, Harenski, & Kiehl, 2013; Decety, Michalska, Akitsuki, & Lahey, 2009; Harenski, Thornton, Harenski, Decety, & Kiehl, 2012; Perino, Moreira, & Telzer, 2019). Further research is needed to examine this possibility.

Concerning the left SI this region has been consistently reported in meta-analyses on motor and tactile imagery (Hardwick, Caspers, Eickhoff, & Swinnen, 2018; Hétu et al., 2013; McNorgan, 2012). Despite that the relationship between retaliatory behaviors and somatosensory cortex is not well understood, the activation of the SI raises the intriguing possibility that subjects may be inclined to mentally simulate aggressive behaviors (e.g., slapping, punching). Indeed, this presumption is supported by studies using script-driven imagery tasks of aggressive behaviors (Herpertz et al., 2017; Treadway et al., 2014). However, it should be noted that the SI is found to be significantly activated in some but not all meta-analyses on pain perception (Duerden & Albanese, 2013; Friebel et al., 2011; Jensen et al., 2016; Lanz et al., 2011; Tanasescu et al., 2016; Xu et al., 2020), indicating that it may be involved in the sensory-discriminative component of pain. Furthermore, it has been shown that the SI may also be involved when imaging oneself being in pain (Decety et al., 2013; Decety & Porges, 2011; Lamm, Decety, & Singer, 2011). These results nevertheless imply that the decision to retaliate involves an element of mentalization. Considering that current theoretical models on aggression have largely ignored the potential role of the primary somatosensory cortex, future studies should thus aim to clarify its precise role in retaliatory behaviors. In addition, we found that the thalamus and the PAG may also be involved retaliation, albeit observed when using a more liberal statistical threshold (p<0.001 uncorrected). More precisely, activation was seen ventral posterolateral part of the thalamus, compared to the medial dorsal cluster observed in frustration. Activation in this particular region sheds a new light on reactive aggression, since evidence indicates strong projections between the ventral posterolateral thalamus and the S1, which form an important hub in nociceptive pain (Ab Aziz & Ahmad, 2006; Vierck, Whitsel, Favorov, Brown, & Tommerdahl, 2013). As mentioned earlier, the PAG plays an important role in pain modulation (Ong et al., 2019; Tanasescu et al., 2016). Thus, activation of the PAG may modulate pain perception (i.e., thalamus, SI) to accurately prepare innate defensive behavior (action preparation: caudate nucleus) against the source of the blocked reward. Indeed, it has been shown that stimulation of the PAG activates defensive behaviors in rodents and cats, namely the fight-or-flight response (Bandler & Depaulis, 1991; Deng, Xiao, & Wang, 2016; Lovick, 1993). Interestingly, according to Panksepp theory (2004; Panksepp & Zellner, 2004), the main hubs of the RAGE system primarily involve the PAG, the amygdala and the hypothalamus. Nearly a century ago, it has been observed that disrupting the diencephalon (hypothalamus and thalamus) and midbrain (PAG) from the cortex (and upper forebrain) produced rage-like behaviors in cats (Bard, 1934). However, it must be acknowledged that we did not observe any activation in the amygdala, which is a core brain region of the threat system. However, given that retaliation is a behavioral response following decisions that were made to punish/harm others in an emotionally intense context, we argue that the amygdala may not be directly involved in action preparation to harm others but may be rather implicated in the affective response to frustrative events, which further initiates defensive behavioral responses (reactive aggression) through PAG activation.

In sum, we found that frustrative non-reward and retaliatory behaviors are both characterized by bilateral FIC and aMCC/pre-SMA, which support Blair (2016)’s assumption on the role of these regions in reactive aggression. It is interesting to note that these brain regions are commonly activated in a wide range of fMRI tasks (Yarkoni, Poldrack, Nichols, Van Essen, & Wager, 2011) such as those involving pain and empathy (Bzdok et al., 2012; Fallon, Roberts, & Stancak, 2020; Fan, Duncan, de Greck, & Northoff, 2011; Lamm et al., 2011). Indeed, both regions are core hubs of the ventral attention network, namely the salience network which subserve stimulus-driven attention (Uddin et al., 2019). Thus, their involvement in frustrative non-reward and retaliation may indicate detection of salient and relevant stimuli in the environment (source of frustration) across these two contexts in order to adequately respond to it. Nevertheless, evidence suggests that lesions and transcranial stimulation of FIC regions are both associated with exaggerated emotional responses (Agustín-Pavón et al., 2012; Gallucci, Riva, Lauro, & Bushman, 2020; Riva et al., 2017; Riva, Romero Lauro, DeWall, Chester, & Bushman, 2015; Shiba, Kim, Santangelo, & Roberts, 2015; Vergallito, Riva, Pisoni, & Lauro, 2018). Taking together, we argue that FIC and aMCC/pre-SMA may be necessary in detecting salient stimuli and reorienting attention toward the source of frustration, further signalling emotional responses (Frustration). Thus, persistent attention towards this stimulus would thus be crucial for evaluating and initiating relevant behavioral responses to the frustrating object (Retaliation).

### 4.2. Neurobiological evidence of the Frustration-Aggression model in clinical populations

Exaggerated responses to frustration are often reported in several psychopathologies that are at risk for reactive aggression such as children and adolescents with high irritability ODD/CD and adult with BPD/ASPD (Bertsch et al., 2020; Blair, 2016; Leibenluft, 2017). Given the limited research on neurobiological correlates of frustration in clinical population, we nonetheless provide evidence that subjects at risk for reactive aggression may exhibit neural deficits within the frustration-aggression neurobiological model.

In response to expected reward that was frustrated, we showed three subsystems that are likely to be interconnected. For instance, as previously shown, deactivation of reward processing may be crucial in our understanding of neural responses to frustration. Interestingly, studies suggest that greater deactivation of the VS during reward omission was observed in adolescents with externalizing symptoms (Bjork et al., 2010) and adults with substance dependent patients which exhibited high levels of impulsivity and neuroticism (Bjork et al., 2008). Moreover, past research demonstrated deficits in expected value sensitivity within the vmPFC (greater deactivation) was observed in children with disruptive behavior disorders (DBDs) compared to typically developing children (White et al., 2013). Evidence also reveals greater vmPFC deactivation when anticipating non-rewards in adults with high psychopathic traits (Bjork, Chen, & Hommer, 2012). However, given the mixed findings regarding deficits in reward processing among CP/ASPD subjects (Byrd, Loeber, & Pardini, 2014; Dugré et al., 2020; Murray, Waller, & Hyde, 2018), deactivation of the reward system during frustration requires further research. Nevertheless, extensive research supports the role of vmPFC in aggressive subjects. In our recent meta-analysis of resting-state connectivity studies, we observed that the vmPFC was a crucial disconnected hub in CP/ASPD subjects, which was also negatively associated with severity antisocial behaviors (Dugré & Potvin, 2021). Furthermore, lesions (or inactivation) of the OFC/vmPFC are frequently linked with greater emotional reactivity and impulsive aggression (Agustín-Pavón et al., 2012; Berlin, Rolls, & Kischka, 2004; H. A. Berlin, Rolls, & Iversen, 2005; Izquierdo, Suda, & Murray, 2005; Kuniishi et al., 2017; Shiba et al., 2015) likely through a disrupted connectivity with the amygdala (Motzkin, Philippi, Wolf, Baskaya, & Koenigs, 2015). Indeed, evidence strongly support the role of deficient vmPFC-amygdala connectivity as a main neurobiological marker of individuals at risk for aggression (Blair, 2008; Blair, 2016; Blair, 2007; Marsh et al., 2011; Marsh et al., 2008; Yoder, Harenski, Kiehl, & Decety, 2015). Given that the deactivation of the vmPFC is frequently observed in patients with exaggerated emotional response (Grupe, Wielgosz, Davidson, & Nitschke, 2016; Milad et al., 2009; VanElzakker, Staples-Bradley, & Shin, 2018), we argue that a greater vmPFC deactivation in response to frustrative events may disinhibit the threat system which give rises to exaggerated emotional arousal, therefore requiring extensive regulation/control (FIC & aMCC/pre-SMA) in healthy controls. In clinical populations at risk for reactive aggression, research indicates deficient regulatory mechanism when processing negative emotions (Coskunpinar, Dir, & Cyders, 2013; Fairchild et al., 2019; Graziano & Garcia, 2016; Hershberger, Um, & Cyders, 2017; Kohls et al., 2020). For instance, adolescents with bipolar disorder exhibit deficits in vmPFC-FIC functional connectivity when processing feedback in a reward-frustrating task (Ross et al., 2020). Among subjects manifesting aggressive & antisocial behaviors, deficits in dACC/aMCC & FIC are frequently reported in meta-analyses of functional and structural neuroimaging studies (Alegria, Radua, & Rubia, 2016; Dugré & Potvin, 2021; Dugré et al., 2020; Poeppl et al., 2019; Rogers & De Brito, 2016). More specifically, a recent meta-analysis demonstrated that the CP/ASPD subjects fail to activate these regions in response to threatening stimuli (Dugré et al., 2020), concurring with a recent study showing that hyperaggressive mice exhibited decreased activation of the ACC during confrontations (van Heukelum et al., 2021). Here, we provided evidence that clinical populations showed a wide range of neural deficits in brain regions involved in the Frustration-Aggression network which may increase their risk for reactive aggression. More precisely, deficits in the vmPFC may be considered as a neurobiological marker of this series of action given its connectivity with multiple brain regions spanning various systems including reward (VS), affective (amygdala, thalamus, PAG) and stimuli-driven attention (aMCC/pre-SMA & FIC). We further suggest that prominent hypofunction of the aMCC/pre-SMA and FIC in clinical populations may yield in persistent emotional response (anger) towards the source of frustration, therefore increasing substantially the risk for retaliatory behaviors.

Finally, it has been proposed that the threat system (i.e., amygdala, PAG, hypothalamus) may be largely implicated in reactive aggression (Bertsch et al., 2020; Blair, 2016; Crowe & Blair, 2008; Gregg & Siegel, 2001; Lickley & Sebastian, 2018; Panksepp, 2004; Panksepp & Zellner, 2004). However, current models of frustration do not take into account the fact that subjects exhibiting antisocial behaviors generally show brain hypo-reactivity to threatening stimuli (Dugré et al., 2020) and are often characterized as being fearless (Barker, Oliver, Viding, Salekin, & Maughan, 2011; Cote, Tremblay, Nagin, Zoccolillo, & Vitaro, 2002; Frogner, Andershed, & Andershed, 2018; Waller & Wagner, 2019). In this sense, there is a hole in the literature examining how individuals at high risk for reactive aggression may perpetrate such behaviors if they are characterized as emotionally hyporeactive (Blair, 2010b; Harenski & Kiehl, 2010). One explanation is that CU traits may mediate relationships between threatening stimuli and amygdala activity (Blair, Leibenluft, & Pine, 2014; Hyde, Shaw, & Hariri, 2013; Viding, Fontaine, & McCrory, 2012). However, in a recent meta-analysis, Dugré and colleagues (2020) found that the severity of antisocial behaviors was negatively associated with the activity in the amygdala (even after controlling for severity of CU traits), whereas no evidence of such relationship was observed with CU traits across whole-brain studies. Indeed, CU traits were only related to the activity of the amygdala in studies using predefined region-of-interest (Dugré et al., 2020). Interestingly, although predominantly associated with increased risk for proactive aggression, psychopathic individuals also exhibit reactive aggression (Cima & Raine, 2009; Fanti, Frick, & Georgiou, 2009; Fite, Stoppelbein, & Greening, 2009; Reidy, Shelley-Tremblay, & Lilienfeld, 2011). Thus, how would they display reactive aggression if they are mainly characterized by emotional underreponsiveness to threat. Therefore, one possibility that could explain such paradoxical results is that CP/ASPD subjects may need more subjectively salient stimuli to enable hyper-activity of the “usually underresponsive” acute threat system. In this sense, the hypo-reactivity observed in CP/ASPD cannot be generalized to every context. Indeed, given that this population is often described as exhibiting selfish behaviors (Krupp, Sewall, Lalumière, Sheriff, & Harris, 2012), we cannot rule out the possibility that CP/ASPD subjects may rather exhibit emotional hyperarousal in some particular contexts such as when the pursuit of their goals has been interrupted (Blair, 2010a). For instance, blocking an expected reward that is subjectively valuable may be more salient than usual emotional stimuli (e.g., facial expression), therefore enhancing the emotional response as well as the risk for reactive aggression. It is thus unequivocal that there is a crucial need to examine this assumption with novel fMRI tasks that specifically target frustration.

## Limitations

The current meta-analysis is the first to examine the neural mechanisms involved in frustration as defined as the omission of an expected reward as well as to investigate the commonalities in the neural processes involved in frustration and retaliatory behaviors. However, some limitations of this meta-analysis should be noted. First, we sought to examine the series of actions underlying the frustration-aggression theory through a multi-domain meta-analysis. Hence, through conjunction analyses, we assumed the temporal sequences between both events, given that it was not possible to test directly this series of actions and how they influence one another. Second, several factors may mediate the relationship between frustrative events and negative affect such as the goal significance and expectations, but also the repeated frustration or sustained effort (Berkowitz, 1989; Breuer & Elson, 2017). Thus, these factors may partially explain why we observed activation in the threat response system (e.g., amygdala, thalamus, PAG) only when we lowered the statistical threshold (p<0.001 uncorrected) (Yu, Mobbs, Seymour, Rowe, & Calder, 2014). Finally, for most studies, retaliatory behaviors were defined by a condition in which subjects decided the severity of the punishment (noise or points removed). Thus, inter-individual differences in retaliatory behaviors remains unknown (e.g., overt/covert, physical/verbal). Future studies may strive to develop functional neuroimaging tasks, with ecological validity, that specifically examine the interactions between frustration and reactive aggression, and how the level of frustration may influence retaliatory behavior. In these investigations, careful attention will need to be paid to the roles of the aINS and the aMCC. At the methodological level, it will be important to select fMRI acquisition parameters optimized to detect activations in brain regions such the amygdala and the PAG.

## Conclusions

In the current study, we sought to examine the neural complexity of frustration as an inhibiting context (e.g., non-reward processing) rather than merely an emotional response. Following the definition provided by the Frustration-Aggression theory (Berkowitz, 1989), we found that withholding an expected reward is mainly characterized by deactivation of the reward system and activation of brain regions involved in the Midcingulo-insular network and acute threat response. Additionally, spatial overlap between frustration and retaliatory behaviors were observed in bilateral FIC and aMCC/pre-SMA, but the latter also involved brain regions subserving goal-directed action preparation and somato-sensory processes. Thus, the current study provides a novel neurobiological framework of frustration-based impulsive aggression based on meta-analytical evidence. Future studies should aim to develop specific fMRI tasks that specifically target frustration-aggression theory to better understand reactive aggression in clinical populations.

## Data Availability

The data that support the findings of this study are available upon reasonable request from the corresponding authors.

## Acknowledgments

JRD is holder of a scholarship from the Fonds de Recherche du Québec en Santé (FRQS). SP is holder of the Eli Lilly Canada Chair on Schizophrenia Research.

## Footnotes

1 In the current meta-analysis, frustration referred to the inhibiting context during which the expected rewarding object was not obtained due to blockage/interference of goal attainment.

